# Accelerated cancer registration from the National Disease Registration Service to support the NHS-Galleri trial

**DOI:** 10.64898/2026.03.24.26345471

**Authors:** Charlotte Eversfield, Natalia Petersen, Rebecca Smittenaar, Wei Liang, Carlos Rocha, Lauren Harrop, Karen Graham, Nathaniel Dilling, Oliver Tulloch, Donna Lloyd, Peter Sasieni, Brian Rous, Martine Bomb, Sean McPhail

## Abstract

**Purpose:** High-quality population-based cancer registration data is crucial for cancer research. We describe the methodology that the National Disease Registration Service (NDRS) established to accelerate provision of cancer registration data in England to support timely analysis of the NHS-Galleri trial (NCT05611632), assessing the clinical utility of a multi-cancer early detection test for population screening.

**Methods:** NDRS established two accelerated cancer registration products for trial participants: ‘Expedited Core’ (providing complete stage data 6 months after diagnosis) and ‘Expedited Comprehensive’ (providing ≥6 months of follow-up clinical pathway data 11 months after diagnosis). NDRS used pre-existing data sources and methodology, plus enhanced dedicated data liaison support. Timeliness and concordance of accelerated data was assessed 6, 11, and ≥19 months after diagnosis.

**Results:** At 6–10 months after diagnosis, most (98.0%) registrations achieved Expedited Core status; at >10 months, all (100%) registrations were Expedited Comprehensive. For fact of malignant cancer and stage III/IV diagnoses, concordance between registrations at 6 and ≥19 months was 96.7% and 95.6%, respectively.

**Conclusions:** NDRS delivered staged cancer registration data 6 months after diagnosis. High concordance at 6, 11, and 19 months provides confidence in the accelerated data. This supports reporting NHS-Galleri findings more promptly than possible via routine data.

**Highlights:**

- NDRS accelerated cancer registration data provision to support a major clinical trial
- Routine cancer registration data is typically available 18–20 months after diagnosis
- Accelerated staged cancer registration data was available 6 months after diagnosis
- Six, 11 & 19-month data concordance was high, indicating good data quality
- This data successfully supported trial analyses but required additional resource

## 1. Background

The National Disease Registration Service (NDRS), part of National Health Service (NHS) England, delivers curated, quality-assured and near-complete data [1] on all malignant and pre-malignant cancer diagnoses in England. Data is collated as the National Cancer Registration Dataset (NCRD) [1,2]. This data supports health service planning and commissioning, and public health and epidemiological research to drive improvements in outcomes for cancer patients.

One such research project is the large, randomised controlled NHS-Galleri trial (NCT05611632), which aims to assess the clinical utility of a blood-based multi-cancer early detection (MCED) test (Galleri^®^; GRAIL, Inc., Menlo Park, CA, USA) for population screening in asymptomatic individuals aged 50–79 years in England [3]. The primary objective of the NHS-Galleri trial is to demonstrate a reduction in the incidence rate of late-stage cancers (stages III or IV) when MCED screening is added to existing NHS cancer screening programmes.

Follow-up of trial participants was conducted using data from routine NHS datasets provided by NDRS, including the NCRD. The detailed data in the NCRD are submitted by healthcare providers across services in NHS Trusts as part of routine, mandated data collection [1]. Typically, NDRS make cancer registration data available approximately 18–20 months following diagnosis; however, prompt evaluation of NHS-Galleri trial objectives required detailed, high-quality cancer diagnosis and staging information substantially earlier than this. Furthermore, given the currently high public health burden of late-stage cancer diagnoses (typically stage III and IV), which generally have worse outcomes than early-stage diagnoses (stage I and II) [4,5], NDRS established an accelerated cancer registration process for NHS-Galleri trial participants. This aimed to support production of timely trial results while maintaining the high data quality necessary to evaluate the clinical utility of MCED screening [3].

Here, we describe the methods NDRS used to accelerate provision of cancer registration data for NHS-Galleri trial participants, and assess the timeliness and quality of these data.

## 2. Methods

### 2.1. Accelerated cancer registration process

#### 2.1.1. Accelerated cancer registration types

NDRS established an accelerated cancer registration process to provide two accelerated cancer registration products to the NHS-Galleri trial team to support trial endpoint analysis. The first product was ‘Expedited Core’, to provide data 6 months after diagnosis, containing information on the cancer diagnosis date, topography, morphology, behaviour, basis of diagnosis, and, crucially, stage (defined in Table A1). The second was ‘Expedited Comprehensive’, to provide cancer registration data 11 months after diagnosis, containing all Expedited Core registration information plus at least 6 months of follow-up clinical pathway (primarily treatment) data. The 6-month period allowed for most patients to have completed NHS diagnostic cancer pathways. As such, stage at diagnosis and any initial treatment procedures were recorded, maximising availability of data needed for a registration. Treatment data was not required to assess NHS-Galleri trial endpoints, but further follow-up ensured the cancer registration remained up to date (e.g., a cancer stage change following surgery).

#### 2.1.2. Data linkage for NHS-Galleri trial participants

The unblinded trial team at Queen Mary University of London uploaded participant cohort data monthly during the study period to a secure NDRS loading portal. A series of automated processing steps then took place in the live cancer registration system (English National Cancer Online Registration Environment, ‘ENCORE’) [1] (Methods A1). Each time the cohort was loaded, and when healthcare data was submitted to NDRS by clinical services, automated processes would identify and link data to NHS-Galleri trial participants, using NHS numbers and cohort identifiers.

Data was exported from ENCORE to monthly static dataset ‘snapshots’, which contained all cancers registered up to that date. Data for NHS-Galleri participants was stored alongside all other cancer registrations in the NCRD monthly snapshots. Thus, participant registrations integrated the same multiple data sources as all other NCRD cancer registrations, as previously described [1]. Data for NHS-Galleri participants was supplied to the trial team for analysis from these snapshots.

Strict rules were followed to uphold patient confidentiality and, as much as possible, NDRS staff were blinded to participant intervention or control arm assignment. NDRS did not process data or provide information on withdrawn participants beyond their withdrawal date. Data acquired between consent and withdrawal was available to the NHS-Galleri trial team, in line with study consent.

#### 2.1.3. Cancer registration officer (CRO) data processing

Every month, two weeks before the NCRD snapshot was taken, CROs processed NHS-Galleri trial participant data according to the earliest date of data received for each participant. A cancer registration was created once data for the month of diagnosis and the subsequent month had been received. Unprocessed data (i.e., that had been patient matched but not previously reviewed by CROs) was reviewed at least every 2 months and at the Expedited Core and Expedited Comprehensive time points.

CROs reviewed every potential cancer registration for trial participants in line with routine registration processes, plus followed additional steps to accelerate processing (Figure A1). CROs initially created a Provisional registration using all available data (though generally initially limited), attempted to resolve conflicts, and flagged key missing data items to the data liaison team. A few registrations were finalised immediately if additional data submissions were not expected, for example for patients who had died.

At 6 months after diagnosis, if a Provisional registration contained the necessary information, it was processed to an Expedited Core registration. At 11 months after diagnosis, and once an Expedited Core registration also included 6 months of clinical pathway follow-up data, it was processed to an Expedited Comprehensive registration. CROs continued adding further data submissions to Expedited Comprehensive registrations, to ensure records remained up to date and accurate. An overview of the accelerated cancer registration timeline is shown in Figure A2.

#### 2.1.4. Data liaison support

NDRS provided enhanced dedicated data liaison resource to support the processing of NHS-Galleri participant data, funded by GRAIL Bio UK, Ltd. (London, UK).

Prior to the launch of NHS-Galleri in 2021, the data liaison team proactively contacted NHS Trusts within the eight Cancer Alliances involved in trial recruitment. This was to promote the need for timely, complete and accurate data submissions and raise awareness of NDRS’ involvement with the trial.

To maximise data quality and completeness, the data liaison team continuously monitored and reviewed registrations with missing or inconsistent data and worked with NHS Trusts to allow completion of required data items, including cancer stage.

NDRS increased dedicated data liaison support for October and November 2023 to maximise completeness and accuracy of NHS-Galleri trial registration data in the December 2023 snapshot to align with trial analysis timelines.

#### 2.1.5. Quality Assurance

Quality assurance checks were embedded at the individual cancer level and for the overall set of registrations. NDRS provided monthly reports on registration data quality and completeness to the NHS-Galleri trial team, and held regular workshops to discuss the accelerated registration process effectiveness. NDRS also offered continuous follow-up and case review of record-level queries from the trial team to improve data quality.

### 2.2. Timeliness, transitions between registration types, and concordance of cancer registration data

These analyses focus on cancer registrations in the December 2023 snapshot for all NHS-Galleri trial participants (i.e., participants with a cancer diagnosis between their date of enrolment and November 2023), for which increased data liaison support was provided. Analysis inclusion and exclusion criteria are defined in Methods A2.

The number of months between the first day of the diagnosis month (as registrations diagnosed between the first and last day of the month were processed simultaneously) and the last day of the snapshot month (data provision to the trial team) was calculated, hereafter referred to as ‘n months after diagnosis’.

For timeliness analysis, all NHS-Galleri trial participant cancer registrations in the December 2023 snapshot were included. For the analysis of transitions between registration types and concordance, registrations were restricted to diagnoses up to the end of July 2023, allowing at least 6 months since diagnosis in the December 2023 snapshot (this restricted snapshot is hereafter termed the ‘6-month snapshot’) (Figure A2).

#### 2.2.1. Timeliness

Data was categorised into time windows from diagnosis for expected accelerated registration timeframes, as defined in section 2.2: <6 months (Provisional); 6–10 months (Expedited Core); 11–18 months (Expedited Comprehensive); and ≥19 months (routine cancer registration).

The percentage of Provisional, Expedited Core and Expedited Comprehensive registrations in the December 2023 snapshot by diagnosis month and year, and in each time window, was calculated to assess registration timeliness.

#### 2.2.2. Cancer registration transitions between registration types

Registrations for diagnoses up to the end of July 2023 were identified in each monthly snapshot up to January 2025. The May 2024 and January 2025 snapshots are hereafter referred to as the 11- and 19-month snapshots, respectively, allowing for at least 11 and 19 months since diagnosis (Figure A2).

The number of Provisional, Expedited Core and Expedited Comprehensive registrations in each snapshot were extracted to assess registration transitions between snapshots.

#### 2.2.3. Concordance

Pairwise comparisons of registrations in the 6-, 11- and 19-month snapshots were made for: fact of malignant cancer; cancer stage; stage III; stage IV; stage III or IV; topography; morphology; and behaviour (Table A1).

Concordance was calculated as the percentage of data that matched out of all cancer registrations present in either comparison snapshot. The denominator for fact of malignant cancer, stage III, stage IV, and stage III or IV cancers, was the number of registrations for which data was recorded in only these categories in either snapshot. Concordance was also calculated by registration type.

The reasons for discordance were extracted where data was inconsistent between snapshots (described in Methods A3).

#### 2.2.4. Statistical analyses

Analyses were performed in R version 4.4.1 (packages detailed in Methods A4). Analytical code is available on GitHub [6].

## 3. Results

### 3.1. Timeliness in the December 2023 snapshot

Of NHS-Galleri trial cancer registrations in the December 2023 snapshot, 2.8% were Provisional, 28.5% Expedited Core, and 68.7% Expedited Comprehensive (Figure 1). The NHS-Galleri trial started enrolling participants on 31st August 2021, and diagnosis dates ranged from October 2021 to November 2023 (Figure 1). Most cancer registrations were Provisional less than 6 months after diagnosis, Expedited Core at 6– 10 months, and all registrations were Expedited Comprehensive after 11 months (Figure 1), in accordance with accelerated registration timeline targets.

**Figure 1.**
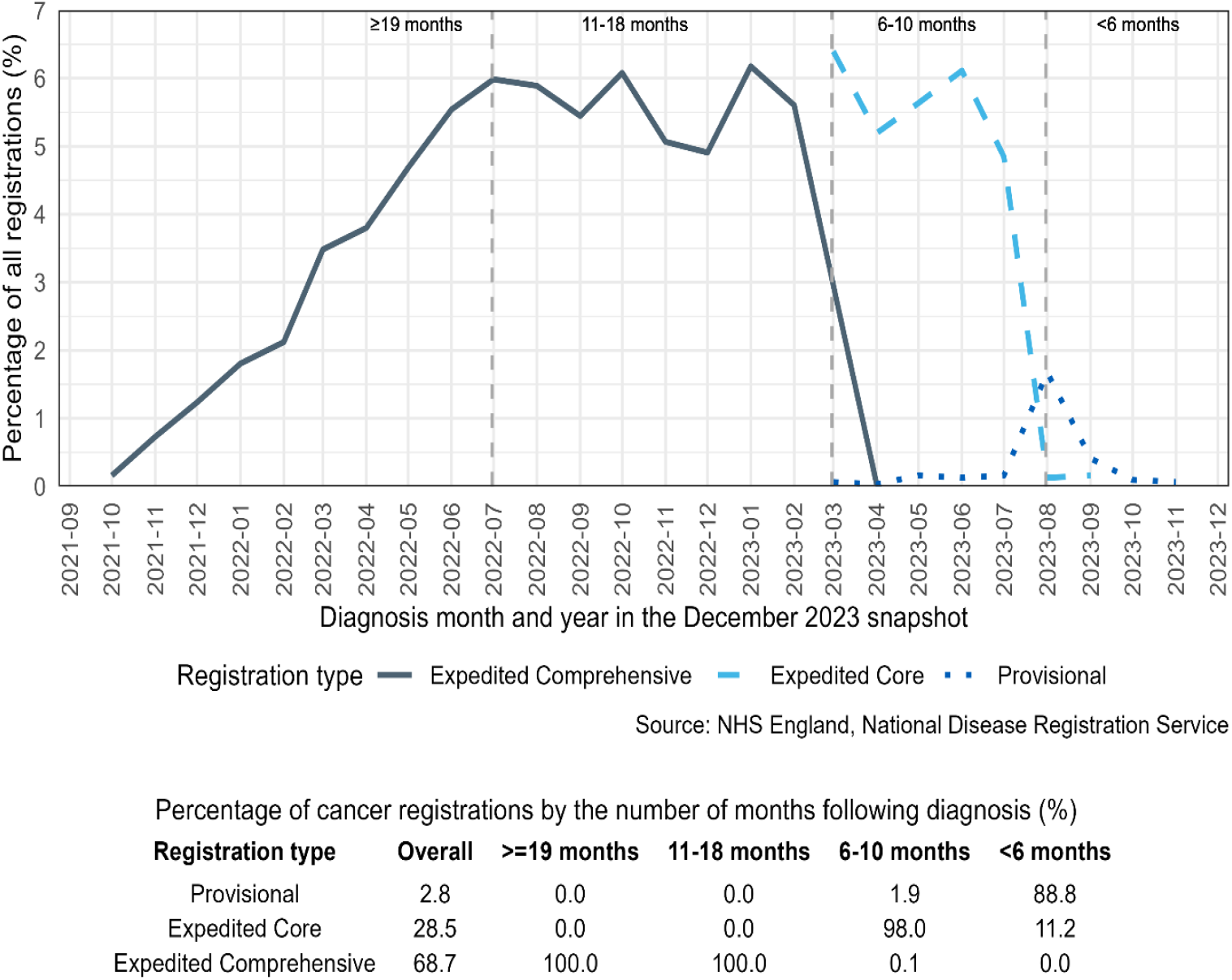
Percentage of NHS-Galleri trial cancer registrations in the December 2023 snapshot by registration type and diagnosis date (month and year), relative to all NHS-Galleri trial cancer registrations in the snapshot. Each frame, separated by the dotted vertical lines, delineate the <6 month (right), 6–10 month (middle right), 11–18 month (middle left), and ≥19 month (left) windows of time between diagnosis and the snapshot date. NHS-Galleri trial participant enrolment began on the 31st August 2021.

### 3.2. Transitions of cancer registration type between snapshots

For registrations in the 6-month snapshot (restricted to diagnoses up to the end of July 2023), 0.55%, 28.91%, and 70.53% were Provisional, Expedited Core, and Expedited Comprehensive, respectively. The percentage of Provisional and Expedited Core registrations decreased over time and in the 19-month snapshot 99.97% of registrations were Expedited Comprehensive, and only 0.03% of registrations were Provisional (Table 1, Figure A3). Most registrations either remained Expedited Comprehensive or progressed from Expedited Core to Expedited Comprehensive between snapshots (Figure 2, Table A2).

**Table 1.** Percentage of NHS-Galleri trial cancer registrations in data snapshots from 6 to 19 months after diagnosis by registration type, for diagnoses up to the end of July 2023. Also displayed as a bar chart in Figure A3.

| Snapshot | Percentage of cancer registrations in each cancer registration type (%) |  |  |
| --- | --- | --- | --- |
|  | Provisional | Expedited Core | Expedited Comprehensive |
| 6 month snapshot | 0.55 | 28.91 | 70.53 |
| 7 month snapshot | 1.00 | 22.74 | 76.26 |
| 8 month snapshot | 0.29 | 18.15 | 81.56 |
| 9 month snapshot | 0.22 | 12.40 | 87.38 |
| 10 month snapshot | 0.16 | 5.91 | 93.93 |
| 11 month snapshot | 0.06 | 0.03 | 99.90 |
| 12 month snapshot | 0.03 | 0.00 | 99.97 |
| 13 month snapshot | 0.00 | 0.00 | 100.00 |
| 14 month snapshot | 0.00 | 0.00 | 100.00 |
| 15 month snapshot | 0.00 | 0.00 | 100.00 |
| 16 month snapshot | 0.03 | 0.00 | 99.97 |
| 17 month snapshot | 0.00 | 0.00 | 100.00 |
| 18 month snapshot | 0.00 | 0.00 | 100.00 |
| 19 month snapshot | 0.03 | 0.00 | 99.97 |

**Figure 2.**
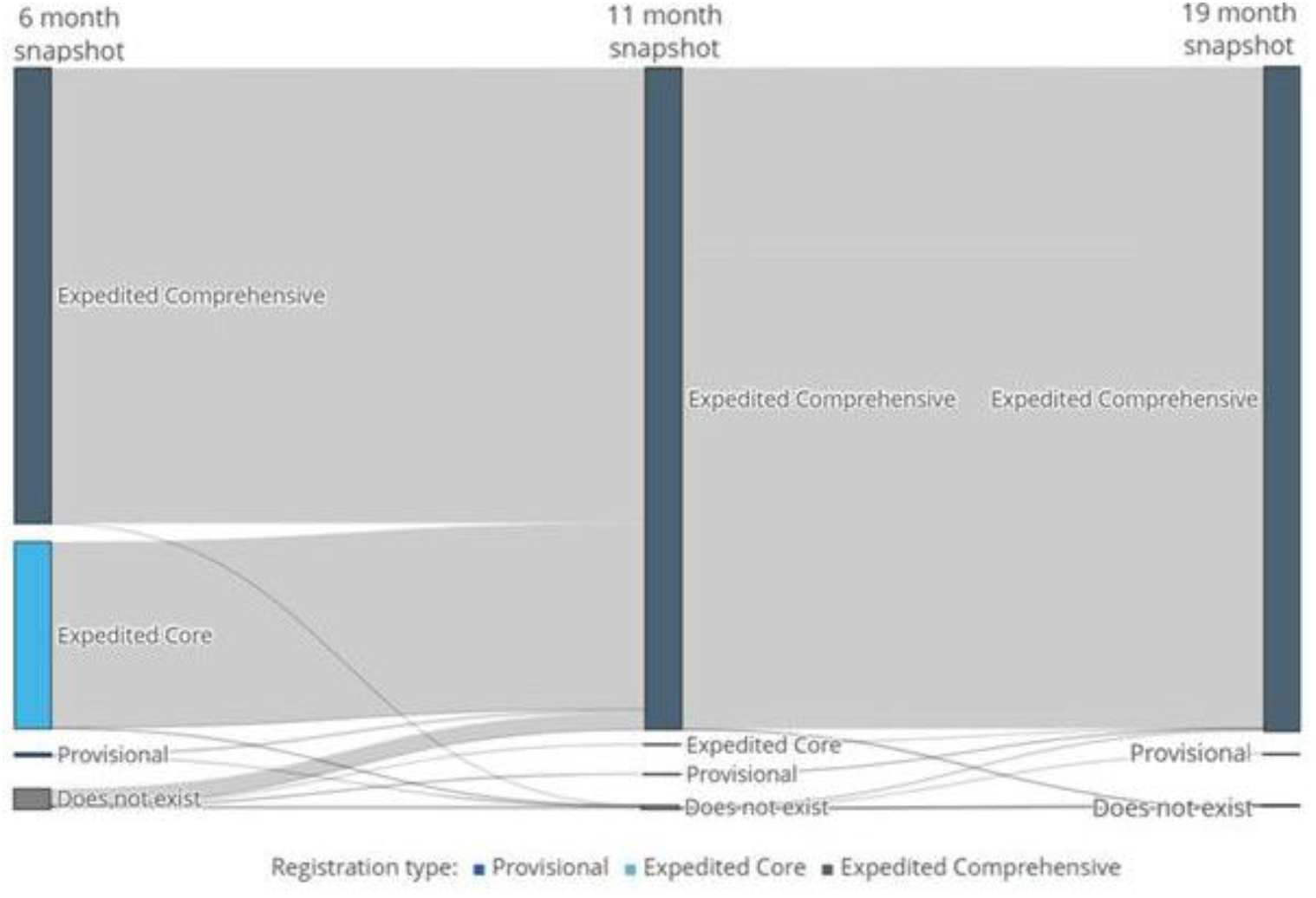
NHS-Galleri trial cancer registrations from the 6-month snapshot to the 11- and 19-month snapshots by registration type (Provisional, Expedited Core, or Expedited Comprehensive) for NHS-Galleri trial participants with diagnoses up to the end of July 2023. Percentages can be seen in Table A2.

In pairwise comparisons of the 6-, 11-, and 19-month snapshots, only a small percentage (≤3.1%) of cancer registrations did not exist in the baseline comparator snapshot, with a small number due to a diagnosis date change to after July 2023, or a behaviour code change (Table 2).

**Table 2.** Registrations observed between NHS-Galleri trial registrations in the 6-, 11-, and 19-month snapshots, for diagnoses up to the end of July 2023.

| Snapshot comparison | Baseline snapshot | Percentage observed in the baseline snapshot only (%) | Reason registrations were only observed in the baseline snapshot (%) |  |  |
| --- | --- | --- | --- | --- | --- |
|  |  |  | Behaviour code change | Diagnosis date after July 2023 | Did not exist in comparison snapshot |
| **6 vs 11 months** | 6 month snapshot | 0.2 | 16.7 | 50.0 | 33.3 |
|  | 11 month snapshot | 2.7 | 4.7 | 29.4 | 65.9 |
| **11 vs 19 months** | 11 month snapshot | 0.1 | 0.0 | 0.0 | 100.0 |
|  | 19 month snapshot | 0.4 | 0.0 | 50.0 | 50.0 |
| **6 vs 19 months** | 6 month snapshot | 0.2 | 14.3 | 42.9 | 42.9 |
|  | 19 month snapshot | 3.1 | 4.1 | 29.6 | 66.3 |

### 3.3. Concordance of cancer registrations between snapshots

Concordance across key variables was high between the snapshots at 6 and 19 months (≥93.7%), 6 and 11 months (≥94.4%), and 11 and 19 months (≥99.1%) after diagnosis (Table 3). Concordance between registrations at 6 and 19 months after diagnosis was 96.7% for fact of malignant cancer and 95.6% for stage III/IV cancer.

**Table 3.** Percentage of NHS-Galleri trial registrations with concordant data between the 6-, 11-, and 19-month snapshots, by variable, and registration type, for diagnoses up to the end of July 2023. Total concordance and total discordance may not sum to 100% due to rounding.

| Snapshot comparison | Cancer registration type comparison | Percentage of concordance for each key variable (%) |  |  |  |  |  |  |  |
| --- | --- | --- | --- | --- | --- | --- | --- | --- | --- |
|  |  | Fact of malignant cancer<sup>a</sup> | Topography | Morphology | Behaviour | Stage III | Stage IV | Stage III or IV | Stage |
| **6 vs 11 months** | All registrations | 97.1 | 96.6 | 96.4 | 97.0 | 94.4 | 97.7 | 96.0 | 95.4 |
|  | Provisional to Expedited Comprehensive | 100.0 | 93.3 | 86.7 | 100.0 | 40.0 | 100.0 | 57.1 | 53.3 |
|  | Expedited Core to Expedited Comprehensive | 100.0 | 98.8 | 98.1 | 99.8 | 89.4 | 97.7 | 93.3 | 95.5 |
|  | Expedited Comprehensive to Expedited comprehensive | 100.0 | 99.9 | 99.8 | 100.0 | 99.8 | 100.0 | 99.9 | 99.7 |
| **11 vs 19 months** | All registrations | 99.5 | 99.4 | 99.1 | 99.5 | 99.2 | 99.4 | 99.5 | 99.1 |
|  | Provisional to Expedited Comprehensive | 100.0 | 100.0 | 100.0 | 100.0 | 0.0 | 0.0 | 0.0 | 50.0 |
|  | Expedited Core to Expedited Comprehensive | 100.0 | 100.0 | 100.0 | 100.0 | 0.0 | 0.0 | 0.0 | 100.0 |
|  | Expedited Comprehensive to Expedited Comprehensive | 100.0 | 99.9 | 99.7 | 100.0 | 99.4 | 99.6 | 99.6 | 99.6 |
| **6 vs 19 months** | All registrations | 96.7 | 96.2 | 95.7 | 96.6 | 93.7 | 97.5 | 95.6 | 94.9 |
|  | Provisional to Expedited Comprehensive | 100.0 | 93.3 | 86.7 | 100.0 | 40.0 | 100.0 | 57.1 | 53.3 |
|  | Expedited Core to Expedited Comprehensive | 100.0 | 98.6 | 97.6 | 99.8 | 88.1 | 97.7 | 92.4 | 95.4 |
|  | Expedited Comprehensive to Expedited Comprehensive | 100.0 | 99.8 | 99.6 | 99.9 | 99.5 | 99.4 | 99.7 | 99.5 |

<sup>a</sup>The denominator differs for all registrations vs registration type comparisons. For all registrations the denominator includes data for which fact of malignant cancer was observed in all registrations for either snapshot; thus the values are not 100%. For registration type comparisons the denominator only includes registrations observed in both snapshots for registration type comparisons; thus concordance is 100%.

Concordance for stage was particularly high for registrations that remained Expedited Comprehensive between snapshots, and for registrations that transitioned from Expedited Core to Expedited Comprehensive, and lowest for registrations that transitioned from Provisional to Expedited Comprehensive (≥99.7%, ≥92.4% and 57.1% respectively for stage III/IV concordance) (Table 3).

Discordance between registrations was mostly due to registrations not existing in the earlier snapshot (Table 4). Most (3.1%) registrations discordant for fact of malignant cancer between the 6- and 19-month snapshots were due to the registration not existing in the 6-month snapshot, compared to 0.2% of registrations not existing in the 19-month snapshot. Some registrations were discordant for stage III/IV cancer, due to the registration having a stage other than III/IV in the earlier snapshot; 1.3% for 6- and 11-months, 0.4% for 11- and 19-months, and 1.6% for 6- and 19-monts.

**Table 4.**
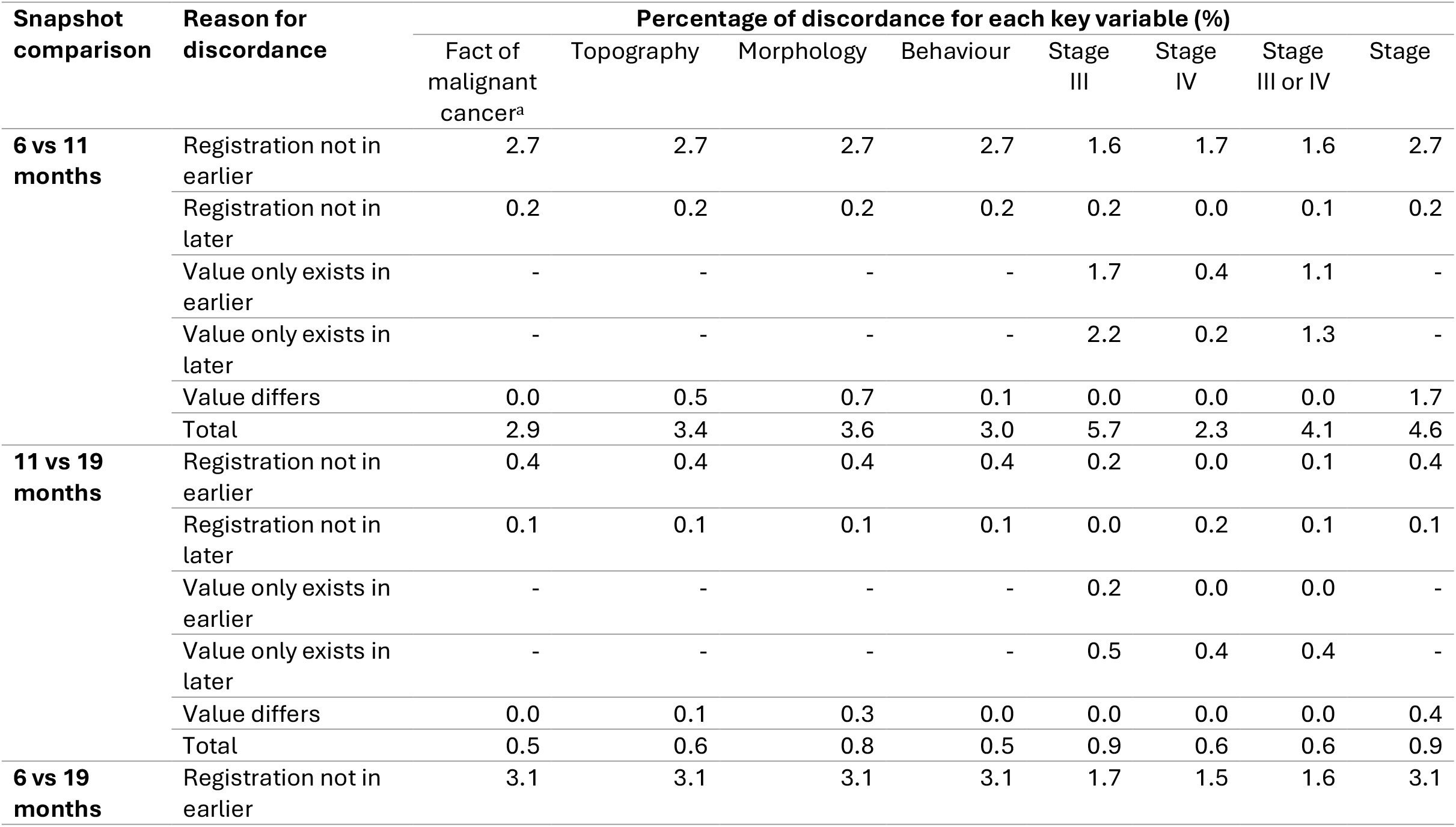
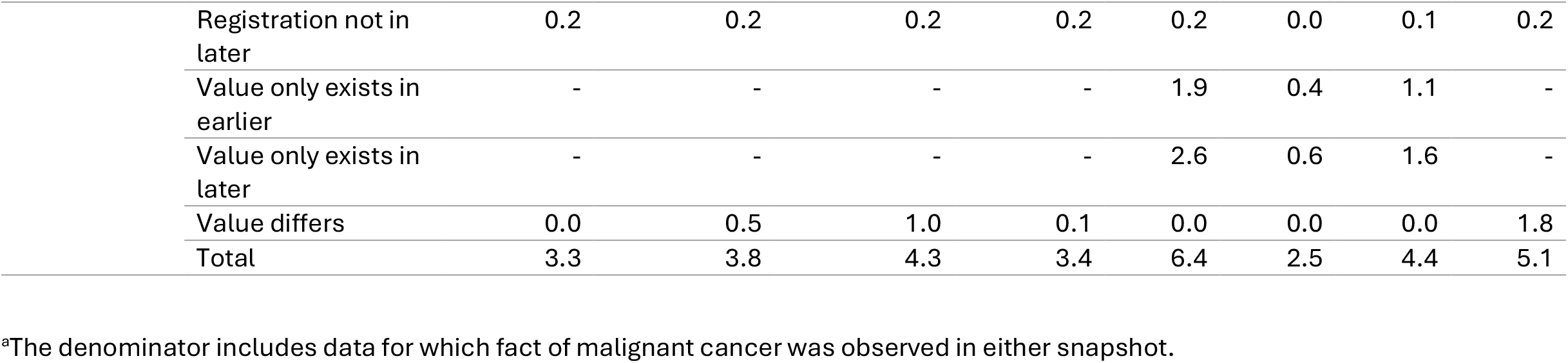
Percentage of NHS-Galleri trial registrations with discordant data between the 6-, 11-, and 19-month snapshots, by variable and reason for discordance, for diagnoses up to the end of July 2023. Total concordance and total discordance may not sum to 100% due to rounding.

## 4. Discussion

NDRS produced accelerated cancer registration data for NHS-Galleri trial participants, with high completeness of key data items, including stage, 6 months after diagnosis. Only a very small percentage of trial participant cancer registrations (1.9%) were Provisional (i.e., missing key data items or required confirmation) 6–10 months after diagnosis. During the NHS-Galleri trial, participants underwent MCED testing annually for three years, with 16–18 months follow-up after the final year of screening; the number of Provisional registrations is therefore expected to decrease with increased follow-up.

Registration data at 6, 11 and 19 months after diagnosis had good concordance for key data items, indicating that the accelerated data were of high quality and accurate, despite being provided substantially earlier than usual NDRS timelines. Discordance was generally due to missing registrations or staging data at earlier time points. NDRS added data to registrations even after the accelerated registration timelines, ensuring that the data is as complete and accurate as possible; hence some discordance is observed for Expedited Comprehensive registrations between snapshots. Only a small percentage of registrations observed in earlier snapshots were not observed in later ones, for example when further information submitted to NDRS indicates that the record represents a condition outside the cohort definition (e.g., a non-malignant neoplasm). Concordance for cancer stage was lowest between Provisional and Expedited Comprehensive registrations as stage data is often incomplete for Provisional registrations, but its presence is a requirement (where a staging system is available for the topography and morphology combination) for progression to Expedited Core or Expedited Comprehensive.

Overall, NDRS’ accelerated timelines were shorter than the median time to cancer registration reported across European cancer registries: between 2010 and 2014, cancer registration data was available at a median of 20 months after diagnosis across 49 European cancer registries [7]. However, a literature search revealed time to cancer registration data are not reported frequently.

The described approach to accelerate registration had some limitations. First, a small percentage of cancer registrations could not be processed within the expected time windows, generally due to: delayed or missing data; data that required further confirmation from providers; delayed responses from providers to data liaison queries; or delays in patient pathways. Whilst it was anticipated that a full 6 or 11 months of follow-up would be required for accurate information, in some scenarios (e.g., patient death or clear documentation of treatment cessation) it was possible to conclude that additional follow-up would be unlikely to alter the data. Nevertheless, there is a risk that registrations submitted early would need to be updated with subsequent additional information, thus decreasing concordance. NDRS produced monthly automated reports monitoring NHS-Galleri registrations, indicating when records were ready for processing and whether they required any further data liaison action. This ensured that issues could be resolved as rapidly as possible during the expected registration time windows. Finally, some data was received from trusts after the expected cancer registration time windows which was still added to the registration to ensure accuracy but may have lowered concordance.

The limitations of this accelerated registration process are balanced by several key strengths. The NCRD is the highest quality dataset available for reliable cancer research and outcomes analysis in England, and is comparable to data collected directly from clinical sites [10]. Curation of data by qualified CROs ensured that cancer registration data made available earlier, for use in the NHS-Galleri trial, were of high quality and consistent with that of routine cancer registration. In addition, NDRS provided ongoing dedicated data liaison support and actively worked with trusts that reported data on NHS-Galleri trial participants to resolve missing or incomplete data, thus enhancing its completeness and accuracy. The increased data liaison support ahead of the December 2023 snapshot further improved the timeliness and quality of registration data for NHS-Galleri participants compared with reports for other months. The same process will be used for the data snapshot which will provide the basis for analysis of the main trial results. Finally, the cancer registration process used already existing cancer registration infrastructure within NDRS, utilising in-house expertise to accelerate cancer registration. Bespoke automated processes were developed to load, prioritise, and monitor the curation of NHS-Galleri trial participant data. These aspects of this novel approach to accelerating cancer registration data were critical to its success.

The NDRS provides high-quality, population-level cancer registration data, incorporating data items from multiple sources, following international best practice [1,11]. As collection of accelerated, high-quality outcome data was an essential component of the NHS-Galleri trial, the trial facilitated additional support to enable the reporting of Expedited Core and Expedited Comprehensive data at 6 and 11 months after diagnosis. This approach required additional upfront and ongoing resources within and in collaboration between various NDRS teams, including the technical infrastructure and development, registration, quality assurance, data liaison, and analytical teams. This enabled delivery of data that is of comparable completeness, concordance and timeliness when compared to cancer diagnosis data collected on clinical trial sites [8]. Using curated data from population-based registries to support the evaluation of clinical trials, that builds on routine flows of healthcare data across the NHS, provides a strong alternative to burdensome and expensive data extraction on clinical trial sites, despite the additional resources needed at the registry to deliver the work. It also enables efficient insights into longer-term follow up, beyond the evaluation of trial primary end points [9].

This analysis demonstrates a successful novel approach to accelerating national registration data for the purposes of a large screening trial. The high concordance of registrations at 6, 11 and 19 months following diagnosis provides confidence in this data to enable trial findings to be reported approximately 13 months sooner than would otherwise be possible. To our knowledge, production of high-quality cancer registration data with high stage completeness at 6 months after diagnosis is a world-first, and underscores the ability of NDRS to successfully adapt routine data flows in the NHS in England, given sufficient resources.

## Data Availability

The data generated in this study are not publicly available to protect the confidentiality of the outcomes data in the ongoing NHS-Galleri trial.

## Abbreviations

CRO: cancer registration officer
ENCORE: English National Cancer Online Registration Environment
MCED: multi cancer early detection
NCRD: National Cancer Registration Dataset
NDRS: National Disease Registration Service
NHS: National Health Service.

## Acknowledgements

This work uses data that has been provided by patients and collected by the NHS as part of their care and support. The data is collated, maintained and quality assured by the National Disease Registration Service, which is part of NHS England. Writing and editorial assistance was provided by Emma B. Saxon (Royston, UK), and was funded by GRAIL Bio UK, Ltd. We would like to thank Andrew Murphy and Christine Head for their support in establishing the accelerated NDRS cancer registration process.

## Author contributions

Conceptualisation: C.E., R.S., W.L., B.R., M.B., S.M.; formal analysis: C.E., N.P., S.M.; investigation: C.E., N.P., R.S., W.L., C.R., L.H., K.G., N.D., D.L., P.S., B.R., M.B., S.M.; methodology: C.E., N.P., R.S., W.L., C.R., L.H., K.G., N.D., D.L., B.R., M.B., S.M.; project administration: C.E., K.G., S.M.; software: O.T.; supervision: K.G., S.M.; validation: C.E., N.P., C.R., L.H., N.D., B.R., M.B.; visualisation: C.E., N.P.; writing – original draft: C.E.; writing – review and editing: C.E., N.P., R.S., W.L., C.R., L.H., K.G., N.D., O.T., D.L., P.S., B.R., M.B., S.M.

