## Supplementary Figure A1 for "Accelerated cancer registration from the National Disease Registration Service to support the NHS-Galleri trial"

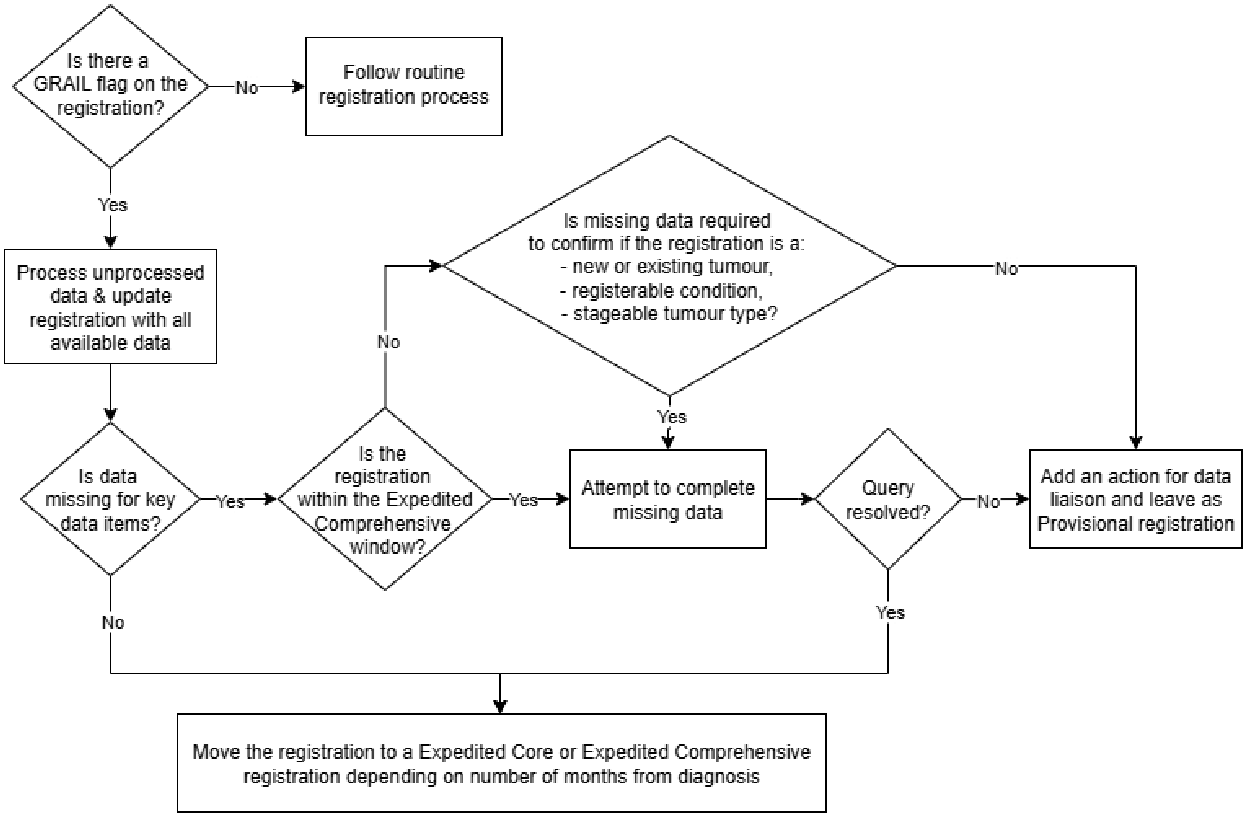


**Figure A1. National Disease Registration Service (NDRS) cancer registration process for registrations with an ‘NHS-Galleri trial participant data’ flag.**
