## Supplementary Figure A2 for "Accelerated cancer registration from the National Disease Registration Service to support the NHS-Galleri trial"

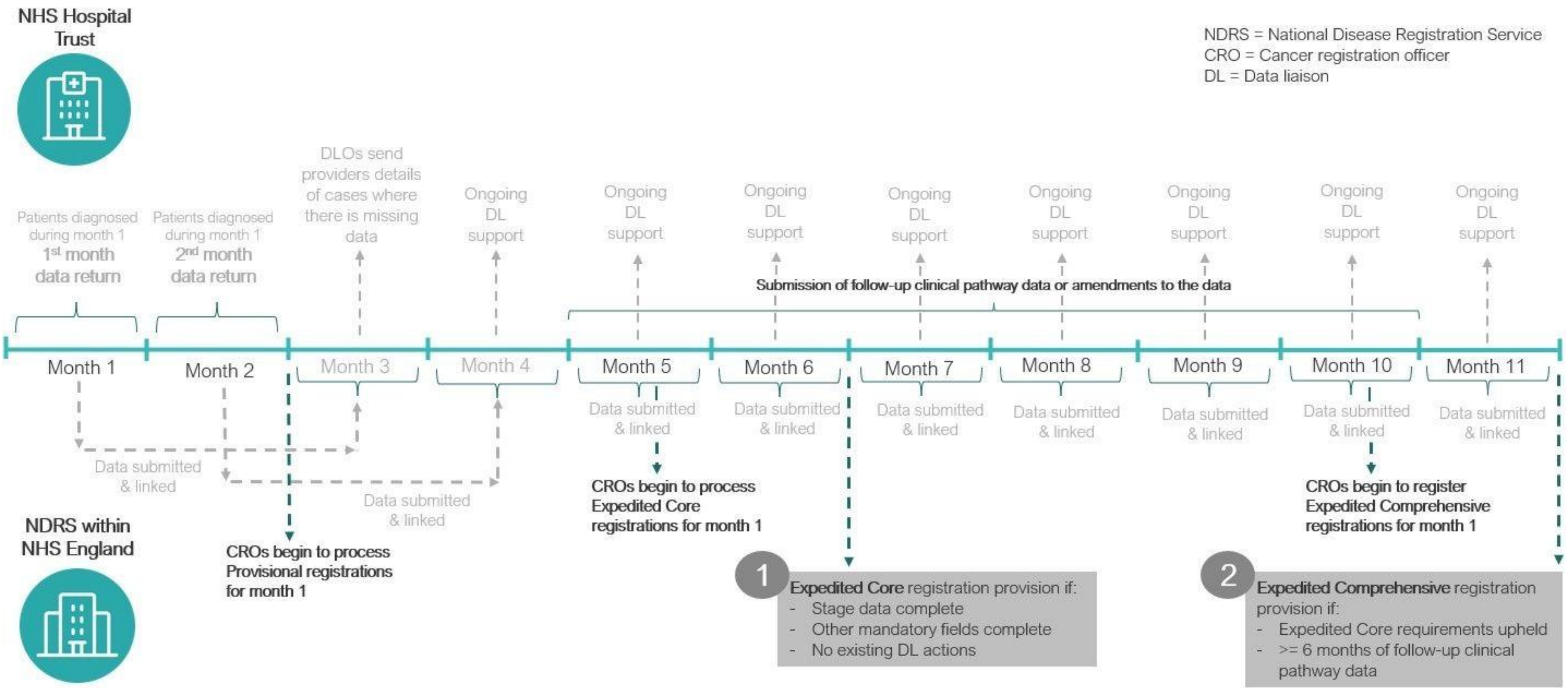


**Figure A2. The National Disease Registration Service (NDRS) accelerated cancer registration timeline for producing Expedited Core and Expedited Comprehensive registrations for NHS-Galleri trial participants, illustrated for diagnoses within 'Month 1'.** As per analyses in this paper, Month 1 corresponds to diagnoses up to the end of July 2023, followed through to Month 6 corresponding to the 6 month snapshot (data from the December 2023 snapshot), and Month 11 corresponding to the 11 month snapshot (data from the May 2024 snapshot).
