## Supplementary Figure A3 for "Accelerated cancer registration from the National Disease Registration Service to support the NHS-Galleri trial"

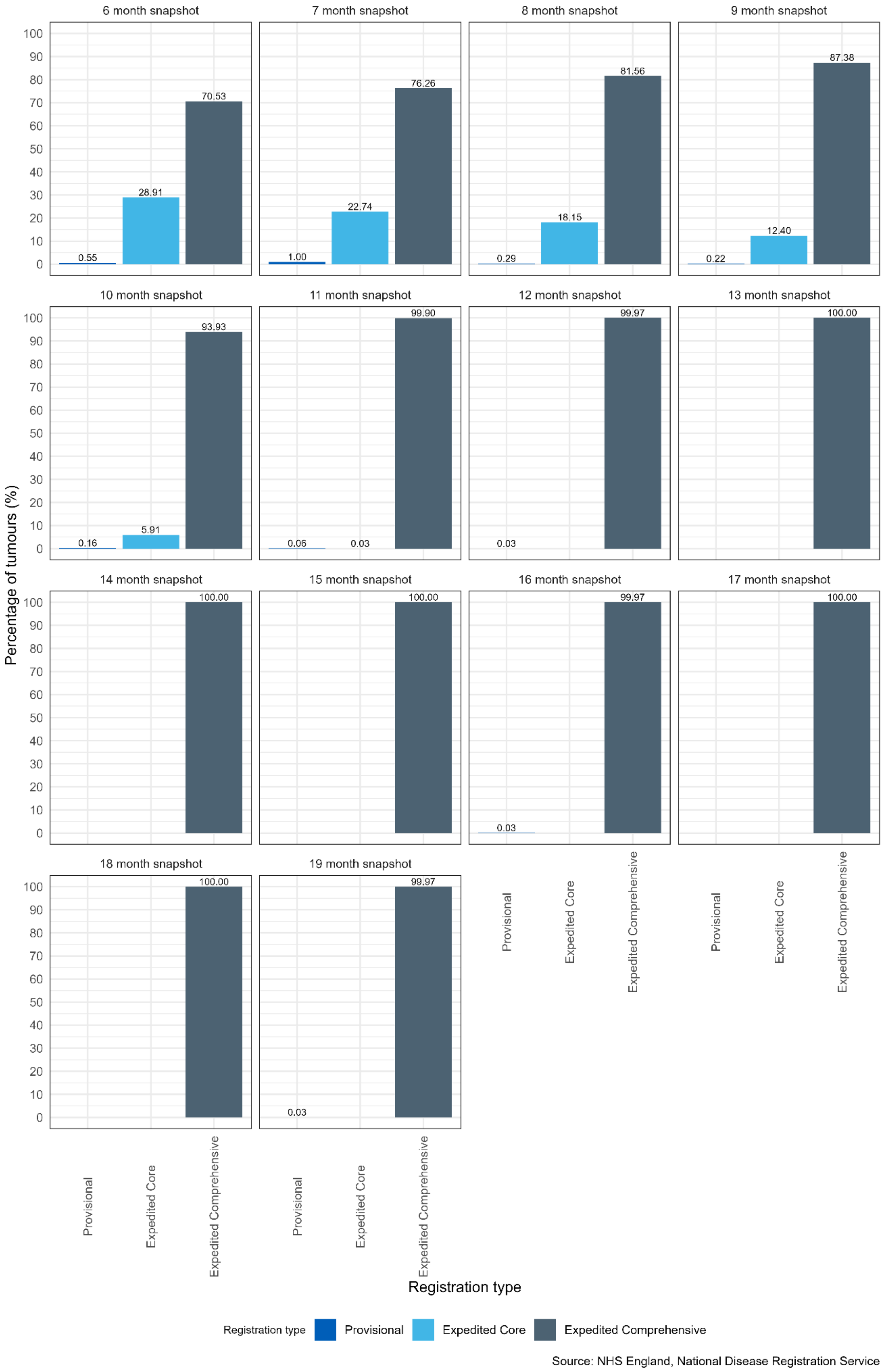

6 month snapshot
7 month snapshot
8 month snapshot
9 month snapshot
10 month snapshot
11 month snapshot
12 month snapshot
13 month snapshot
14 month snapshot
15 month snapshot
16 month snapshot
17 month snapshot
18 month snapshot
19 month snapshot
0.55
28.91
70.53
1.00
22.74
76.26
0.29
18.15
81.56
0.22
12.40
87.38
0.16
5.91
93.93
0.06
0.03
99.90
0.03
99.97
100.00
100.00
100.00
0.03
99.97
100.00
100.00
0.03
99.97
Percentage of tumours (%)
Registration type
Provisional
Expedited Core
Expedited Comprehensive
Registration type
Provisional
Expedited Core
Expedited Comprehensive
Source: NHS England, National Disease Registration Service

**Figure A3. Percentage of NHS-Galleri trial cancer registrations in the snapshots 6 to 19 months after diagnosis by registration type (Provisional, Expedited Core, or Expedited Comprehensive), for diagnoses up to the end of July 2023.**
