## Supplementary Methods A1 for "Accelerated cancer registration from the National Disease Registration Service to support the NHS-Galleri trial"

**Methods A1. Automated processing steps used to produce accelerated cancer registration data for NHS-Galleri trial participants, which take place upon cohort load.**

1. Validation checks on data items in the cohort file, e.g., for NHS number, date formatting, and dates in the future.
2. Disparity checks for participant and patient IDs to check if a participant record has already been submitted.
    a. If so, the record was linked to the existing patient ID, and checks were run for any changes to personal information and demographics, including name, address, gender, and date of birth.
    b. If not, a patient ID was created for the participant, including for those without a cancer diagnosis.
3. Tracing of NHS-Galleri participants against information held by the Personal Demographic Service in NHS Spine, to validate demographics and record vital status information.
4. Generation of automated reports for monitoring NHS-Galleri trial participants, including the number of registrations, whether records were ready for processing, and any withdrawn participants.
