## Supplementary Methods A2 for "Accelerated cancer registration from the National Disease Registration Service to support the NHS-Galleri trial"

**Methods A2. Inclusion criteria used for data linkage and analysis of timeliness, transitions and concordance of cancer registration data for the NHS-Galleri trial.**

1. Participants had not withdrawn from the trial, or the date of their withdrawal was after the date of their diagnosis.
2. Participant age at diagnosis was between 0 and 200 years (this was in order to capture all trial participants; those who were not within the NHS-Galleri trial criterion for age, 50–79 years, were retrospectively considered ineligible by the trial team).
3. Participant recorded gender at diagnosis was 1 (male) or 2 (female).
4. Participant was resident in England at the time of their diagnosis.
5. Cancer registration was confirmed as a non-duplicate registration.
6. Diagnosis date was after or on the same day as enrolment into the trial.
7. Cancer registration had a behaviour code of 3, 5, 6 or 9.
8. Cancer registrations were not basal cell carcinoma or squamous cell carcinoma of the skin, defined as an International Classification of Diseases for Oncology (ICD-O-3) [1] code of C44, C000, C001 or C002, with an ICD-O-3 morphology code between 8051 and 8081 or between 8083 and 8098.
