## Supplementary Methods A3 for "Accelerated cancer registration from the National Disease Registration Service to support the NHS-Galleri trial"

**Methods A3. Reasons for discordance where data was inconsistent between data snapshots (as used in the concordance analysis).**

1. Registration was not in one of the snapshots being compared (either the earlier or later snapshot).
2. Registration was in both snapshots being compared, but was a malignant cancer, or stage III, stage IV, or stage III or IV in only one of the snapshots (either the earlier or the later snapshot), i.e., the value only existed in the earlier or later snapshot.
3. Values for the variable differed between the snapshots e.g. data existed for the variable in both snapshots, but the data in the earlier or later snapshot was different.
