## Supplementary Methods A4 for "Accelerated cancer registration from the National Disease Registration Service to support the NHS-Galleri trial"

**Methods A4. R packages used for analysis**

The following R packages were used for analysis: data.table_1.17.6, dplyr_1.1.4, DT_0.33, ggplot2_3.5.2, here_1.0.1, htmltools_0.5.8.1, htmlwidgets_1.6.4, knitr_1.50, lubridate_1.9.4, NDRSAfunctions_1.0.0, NHSRtheme_0.1.0, pacman_0.5.1, plotly_4.10.4, rJava_1.0-11, stringr_1.5.1, tibble_3.3.0, tidyr_1.3.1, zoo_1.8-14.
