## Supplementary Table A1 for "Accelerated cancer registration from the National Disease Registration Service to support the NHS-Galleri trial"

**Table A1. Definitions of fact of malignant cancer, cancer stage, stage III cancer, stage IV cancer, topography, morphology, and behaviour (as used in the concordance analysis), and basis of diagnosis and date of diagnosis.**

| Key variable | Definition |
|---|---|
| **Fact of malignant cancer** | Any cancer with a behaviour code of 3, 5, 6 or 9. [1] |
| **Cancer stage** | Stage at diagnosis of the tumour determined from data curated by NDRS. |
| **Stage III cancer** | Any cancer with stage III at diagnosis (to 1 character), as recorded in one of the following staging systems: UICC, FIGO, Ann Arbor, R-ISS or ENETS. |
| **Stage IV cancer** | Any cancer with stage IV at diagnosis (to 1 character), as recorded in one of the following staging systems: UICC, FIGO, Ann Arbor, R-ISS or ENETS. |
| **Topography** | Full ICD-O-3 topography code to 4 characters (e.g., C18.1) [1]. |
| **Morphology** | ICD-O-3 cell type (histology) (first four characters) [1]. |
| **Behaviour** | ICD-O-3 behaviour (5th digit code) [1]. |
| **Date of diagnosis** | Conforms with the international requirements specified by the European Network of Cancer Registries (ENCR) for incidence date [2]. |
| **Basis of diagnosis** | The method used to confirm the cancer and to provide a level of certainty of the diagnosis of cancer. Conforms with ENCR recommendations [3]. |
