## Supplementary Table A2 for "Accelerated cancer registration from the National Disease Registration Service to support the NHS-Galleri trial"

**Table A2. Percentage of NHS-Galleri trial registrations by registration type transition through 6, 11, and 19 month snapshots, for diagnoses up to the end of July 2023.**

| Snapshot | | | Percentage of registrations (%) |
|---|---|---|---|
| **6 month snapshot** | **11 month snapshot** | **19 month snapshot** | |
| Provisional | Expedited Comprehensive | Expedited Comprehensive | 0.47 |
| Provisional | Does not exist | Does not exist | 0.06 |
| Expedited Core | Expedited Comprehensive | Expedited Comprehensive | 27.89 |
| Expedited Core | Expedited Comprehensive | Does not exist | 0.03 |
| Expedited Core | Does not exist | Does not exist | 0.09 |
| Expedited Comprehensive | Expedited Comprehensive | Expedited Comprehensive | 68.30 |
| Expedited Comprehensive | Does not exist | Does not exist | 0.03 |
| Does not exist | Provisional | Expedited Comprehensive | 0.06 |
| Does not exist | Expedited Core | Expedited Comprehensive | 0.03 |
| Does not exist | Expedited Comprehensive | Expedited Comprehensive | 2.55 |
| Does not exist | Expedited Comprehensive | Does not exist | 0.03 |
| Does not exist | Does not exist | Provisional | 0.03 |
| Does not exist | Does not exist | Expedited Comprehensive | 0.41 |
